# Assessing the Impact of COVID-19 on Multimorbidity: Insights from Structural Equation Modeling in Colombia

**DOI:** 10.1101/2024.09.11.24313500

**Authors:** A Porras-Ramírez, A Rico-Mendoza, A Campos-Maya, J Cardenas-Sanchez, D Penagos-Lopez, A Gomez-Puentes, N Delgado-Quiroz, L Guarin-Muñoz, J Ortiz-Elejalde

## Abstract

**Background:** Multimorbidity, the coexistence of multiple chronic diseases, poses significant challenges for healthcare systems worldwide. This study aims to assess the situation of multimorbidity in Colombia using structural equation models and to characterize multimorbidity by sex, age group, region, and health insurance regime between 2019 and 2023.

**Methods:** We conducted a cross-sectional analytical study utilizing data from the Individual Health Service Provision Registry (RIPS) and other national databases, including the Unit of Payment by Capitation (UPC) Sufficiency Study Database. Structural equation models were employed to identify and analyze multimorbidity clusters among patients with two or more chronic diseases.

**Results:** The study revealed a multimorbidity prevalence of 61.5% in the general population. Hypertension emerged as the most prevalent chronic condition, frequently associated with diabetes and chronic kidney disease. Multimorbidity was more common in women and individuals affiliated with the contributory insurance regime. The study also identified significant variations in multimorbidity prevalence across different regions and age groups.

**Conclusions:** Multimorbidity is a prevalent issue in Colombia, with substantial variations by sex, age, and insurance regime. These findings underscore the need for tailored healthcare strategies to address the diverse needs of multimorbid patients. The results provide critical insights for health service planning and management, emphasizing the importance of continued research and policy development to mitigate the burden of chronic diseases and multimorbidity in Colombia.

## Background

Structural equation models (SEMs) are advanced analytical tools that estimate the effects and relationships between multiple variables, offering greater flexibility than traditional regression models (1). Combining elements of factor analysis and path analysis, SEMs model both direct and indirect effects among variables, making them particularly valuable in health research where complex interrelationships are common (2). A key strength of SEMs is their ability to construct latent variables, enhancing the reliability and validity of the models by capturing underlying constructs that traditional regression models might miss (Byrne, 2010). SEMs also allow for the simultaneous estimation of multiple relationships, providing a comprehensive understanding of the data.

In recent years, the prevalence and burden of multimorbidity—the presence of two or more chronic conditions—have gained increasing attention globally. Multimorbidity poses significant challenges for healthcare systems in terms of healthcare planning, resource allocation, and patient management (3). Understanding the distribution and determinants of multimorbidity in Colombia is critical for effective health policy and intervention design.

This study aims to assess multimorbidity in Colombia by characterizing the multimorbidity patterns of individuals affiliated with the General System of Social Security in Health (SGSSS) from 2019 to 2023. Utilizing SEMs to analyze multimorbidity clusters, the study examines variations by sex, age group, region, and insurance regime, providing insights into the complex interactions of chronic diseases and informing health service planning and policy development in Colombia (4–6).

### Colombian Health System and Affiliation Regimes

Colombia’s health system, structured under the General System of Social Security in Health (SGSSS) established by Law 100 of 1993, aims to provide universal health coverage to all Colombians. The SGSSS operates through two main insurance regimes: the contributory regime and the subsidized regime. This dual-regime structure promotes equity in healthcare access by offering different pathways to health insurance based on financial ability, ensuring that all citizens, regardless of economic status, can receive necessary medical care. This approach helps reduce health disparities and improve overall public health in the country.

### Prevalence of Hypertension and Diabetes and healthcare utilization

Before 2012-2019, hypertension and diabetes prevalence in Colombia was stable at 20.5%, indicating consistent chronic disease management and healthcare access. During COVID-19, prevalence surged to 24.3% in 2021 due to severe COVID-19 risks, increased diagnosis, and healthcare disruptions worsening chronic conditions.

SEM analysis showed that prior to the pandemic, patients with hypertension and diabetes had 6.8 healthcare visits annually and a 78% medication adherence rate. During COVID-19, visits decreased to 5.4 in 2020, with adherence at 65%, due to infection fears and healthcare strain. By 2023, visits increased to 6.2 yearly, adherence improved to 78%, aided by healthcare adaptations and telemedicine, as restrictions eased.

### Hospitalization Rates

Before the COVID-19 pandemic (2012-2019), hospitalization rates for hypertension and diabetes complications were around 25 per 1000 patients. This rate increased to 38 per 1000 in 2020 due to the severe impact of COVID-19 and delayed care. By 2023, rates declined to 30 per 1000, indicating improvements in chronic disease management, healthcare access, vaccination efforts, and effective COVID-19 treatments.

## Methods

### Study Design

This study is an exploratory ecological investigation aimed at defining multimorbidity clusters in Colombia and characterizing these clusters according to age group, sex, region, and affiliation with the General System of Social Security in Health (SGSSS).

### Study Period

The analysis covers data collected from 2019 to 2023.

### Data Sources

The primary sources of data for this study include:

1. Individual Health Service Provision Registry (RIPS): This registry contains detailed records of health services provided, including identification of health service providers, users receiving the services, types of services provided, and diagnoses. The RIPS database categorizes data into identification data, health service data, and reason for service data.
2. Unit of Payment by Capitation (UPC) Sufficiency Study Database: This database contains EPS-provided data, quality-checked by the Ministry of Health and Social Protection. It covers healthcare expenses and technologies categorized into activities, interventions, procedures, medications, devices, and supplies.
3. Additional Databases:

- Vital Statistics Database from DANE
- Unique Affiliate Database (BDUA)
- Reimbursement Database
- MIPRES Database
- High-Cost Account Database
- SIVIGILA Database

### Population and Sample

The study population consists of individuals affiliated with the contributory and subsidized regimes of SGSSS during the study period. The units of observation are patients with two or more chronic diseases reported in the sufficiency and RIPS databases (7).

### Variables

Key variables include age, sex, department, prevalence of chronic conditions, disability status, and health service usage. The presence of chronic conditions is treated as a binary variable in the structural equation model.

### Analytical Methods

1. Systematic Literature Review: A comprehensive literature review was conducted to establish definitions of interest, identify clusters from multimorbidity studies in other countries, and determine the methodology for identifying and prioritizing disease clusters.
2. Selection and Grouping of ICD-10 Diagnoses: Diagnoses from the International Classification of Diseases 10th Revision (ICD-10) were classified to identify chronic diseases. An agreement exercise between the World Bank and Ministry of Health and Social Protection teams facilitated the grouping of diseases to identify multimorbidity clusters.
3. Age Group Definition: The population was divided into the following age groups for analysis:

- 0 to 17 years: Childhood and Adolescence
- 18 to 59 years: Adulthood
- 60 years and older: Elderly
4. Structural Equation Modeling (SEM): SEM calculated crude prevalence of global and stratified multimorbidity by age and sex. It estimated causal relationships from statistical data and qualitative assumptions, calculating frequencies of all possible multimorbidity patterns. Significance was set at α = 0.005, analyzed using Stata Statistical Software (version 14).
5. Factor Analysis: Factor analysis was employed to reduce and summarize the data, calculating latent dimensions (factors) to explain relationships among variables. The simplest factorial structure was extracted for easier interpretation.

### Statistical Analysis

The SEM was validated against empirical data samples to determine if the data fit the hypothesized model, with statistical significance assessed using a chi-square test (p-value of X² = 0.0023). Factor analysis was used to identify and interpret underlying dimensions of multimorbidity.

This methodology ensures a comprehensive and rigorous approach to understanding multimorbidity in the Colombian population, providing valuable insights for healthcare policy and management.

## Results

For this study, a structural equation was designed considering variables such as age, sex, department, prevalence, disability, and service usage to identify and prioritize the main clusters of chronic diseases affecting health service delivery during the analysis period.

Data for the structural equation model came from insurers of Colombia’s contributory and subsidized regimes, using the SGSSS sufficiency study database from 2019 to 2023. It covered the general population, segmented by age groups (0-17, 18-59, and 60+), sex, insurance regime, and department. Coverage details by insurance regime are provided in Table 1.

**Table 1.** Coverage by Insurance Regime in the Sufficiency Data. (**2019–2023**)

| Regime | 2019 | 2020 | 2021 | 2022 | 2023 |
| --- | --- | --- | --- | --- | --- |
| Contributory | 19,957,738 | 20,150,266 | 20,750,202 | 21,295,724 | 21,713,023 |
| Subsidized | 22,605,295 | 22,669,543 | 22,596,377 | 22,016,310 | 21,176,856 |
| Sufficiency (C) | 14,041,322 | 15,153,132 | 15,936,771 | 16,550,231 | 16,801,472 |
| Sufficiency (S) | 8,694,001 | 8,919,836 | 8,908,969 | 10,971,130 | 11,802,194 |
| Coverage (C) % | 70.4 | 75.2 | 76.8 | 77.7 | 77.4 |
| Coverage (S) % | 38.5 | 39.3 | 39.4 | 49.8 | 55.7 |

### Proportion of Patients with Multimorbidity

From 2019 to 2023, multimorbidity affected 61.5% of the general population, amounting to an annual average of 11,192,469 individuals (8). The distribution of conditions showed 29.1% with two conditions, 11.4% with three, and decreasing percentages with higher numbers of conditions (Table 2).

**Table 2.** Proportion of Patients with Multimorbidity Using Structural Equation in Colombia (2019–2023)

| Year | 1 Condition | 2 Conditions | 3 Conditions | 4 Conditions | 5 Conditions | 6 Conditions | 7 Conditions | Total Conditions |
| --- | --- | --- | --- | --- | --- | --- | --- | --- |
| 2019 | 5,132,523 | 2,875,456 | 1,075,517 | 544,375 | 136,521 | 93,253 | 78,520 | 9,936,165 |
| % | 51.7 | 28.9 | 10.8 | 5.5 | 1.4 | 0.9 | 0.8 | 1000 |
| 2013 | 5,118,925 | 2,928,027 | 1,096,053 | 553,536 | 142,160 | 96,663 | 80,887 | 10,016,251 |
| % | 51.1 | 29.2 | 10.9 | 5.5 | 1.4 | 1.0 | 0.8 | 1000 |
| 2014 | 6,076,510 | 3,402,977 | 1,337,729 | 675,290 | 204,892 | 128,601 | 96,932 | 11,922,931 |
| % | 51.0 | 28.5 | 11.2 | 5.7 | 1.7 | 1.1 | 0.8 | 1000 |
| 2015 | 6,102,653 | 3,455,548 | 1,358,265 | 684,451 | 210,531 | 132,011 | 99,299 | 12,042,758 |
| % | 50.7 | 28.7 | 11.3 | 5.7 | 1.7 | 1.1 | 0.8 | 1000 |
| 2023 | 6,128,796 | 3,508,119 | 1,378,801 | 693,612 | 216,170 | 135,421 | 101,666 | 12,044,240 |
| % | 50.9 | 29.1 | 11.4 | 5.8 | 1.8 | 1.1 | 0.8 | 1000 |

Hypertension was the most prevalent condition across all age groups in the main clusters identified, with an overall prevalence of 41.2% in the general population, increasing with age. The table was constructed using a probability threshold of 0.1, indicating all clusters listed exceeded this probability.

### Distribution of Multimorbidity by Age

For the analyzed age groups, it was found that in the 0 to 17 years group, chronic periodontal diseases were most common (65.3%), with multimorbidity at 15.3% annually (838,530 individuals). Common clusters included vision disorders and periodontal diseases (probability 0.87). Among those aged 18-59, multimorbidity was 33.1% (1,814,074 individuals), with chronic back pain prevalent (65.0%). Common clusters included chronic back pain and periodontal diseases (probability 0.90). In adults over 60, multimorbidity was 51.3% (2,811,541 individuals), with hypertension most prevalent (68.7%). Common clusters included hypertension and diabetes (probability 0.96), and hypertension, diabetes, and chronic kidney disease (0.91). Fifty clusters were identified using a probability cutoff of 0.1.

### Distribution of Multimorbidity by Sex and SGSSS Affiliation

From 2019 to 2023, multimorbidity was more prevalent among women (67.3%) and was higher in the contributory insurance regime (58.3%). This reflects greater healthcare utilization by contributory regime patients, who are more represented in the UPC sufficiency database compared to subsidized regime patients (9).

### Distribution of Multimorbidity by Department

The highest prevalence of multimorbidity by department was found in Risaralda, Quindío, Huila, and Magdalena (prevalence proportion higher than 7,000 per 100,000 inhabitants) (Map 1).

**Map 1.**
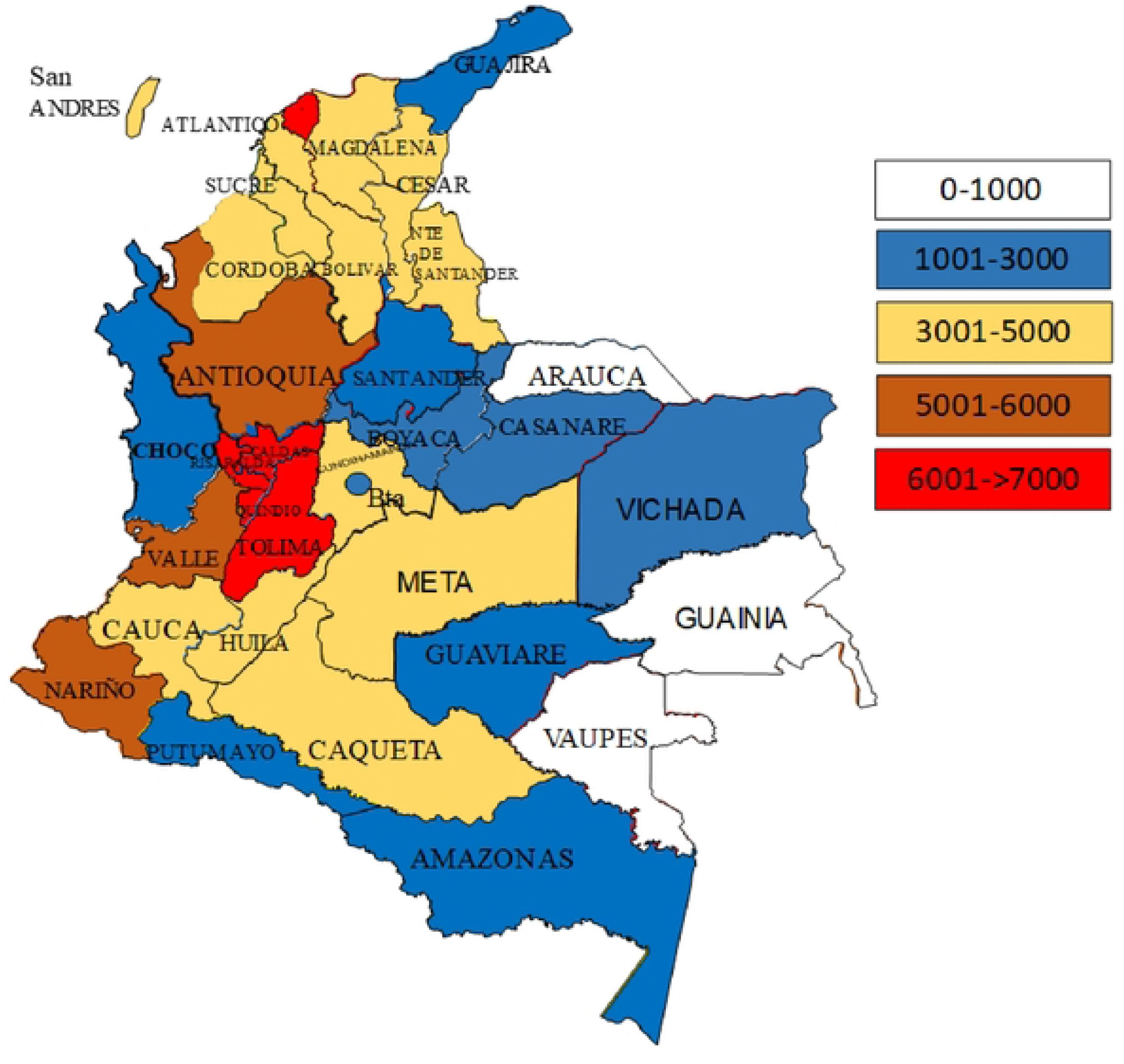
Proportion of Multimorbidity Prevalence by Department in Colombia (2019-2023)

Map 1. Proportion of Multimorbidity Prevalence by Department in Colombia (2019-2023)

The main clusters of multimorbidity by department are shown in Table 3.

**Table 3.** Most Frequent Clusters by Department in Colombia (2019-2023)

| Department | Most Frequent Clusters | Probability |
| --- | --- | --- |
| Antioquia | Hypertension and Diabetes | 0.82 |
|  | Prostate Cancer and Hypertension | 0.74 |
|  | Hypertension and Chronic Kidney Disease | 0.72 |
|  | Hypertension, Diabetes, and Chronic Kidney Disease | 0.71 |
|  | Chronic Viral Hepatitis and Malignant Liver Tumors | 0.51 |
|  | Malignant Breast Tumor, Obesity, and Hypertension | 0.51 |
|  | Chronic Viral Hepatitis and Malignant Liver Tumors | 0.50 |
|  | Leukaemia and Substance Use Disorders | 0.49 |
| Atlántico | Hypertension and Chronic Kidney Disease | 0.70 |
|  | Hypertension and Diabetes | 0.65 |
|  | Thyroid Cancer, Obesity, and Diabetes | 0.65 |
|  | Hypertension, Diabetes, and Chronic Kidney Disease | 0.62 |
| Bogotá | Prostate Cancer and Hypertension | 0.74 |
|  | Prostate Cancer and Hypertension | 0.74 |
|  | HIV/AIDS and Tuberculosis | 0.64 |
|  | HIV/AIDS, Tuberculosis, and Sequelae | 0.64 |
|  | Colon Cancer and Hypertension | 0.62 |
|  | Cervical Cancer and Depression | 0.52 |
|  | Lymphoma and Hypertension | 0.52 |
|  | Hypertension and Lymphoma | 0.52 |
| Bolívar | Hypertension and Chronic Kidney Disease | 0.78 |
|  | Hypertension and Diabetes | 0.71 |
|  | Hypertension, Diabetes, and Chronic Kidney Disease | 0.65 |
|  | Hypertension and Cerebrovascular Diseases | 0.61 |
| Boyacá | Hypertension and COPD | 0.66 |
|  | Hypertension and Malignant Stomach Tumors | 0.64 |
|  | Hypertension, Diabetes, and Chronic Kidney Disease | 0.63 |
|  | Hypertension and Chronic Kidney Disease | 0.62 |
|  | Hypertension and Diabetes | 0.60 |
| Caldas | Hypertension and Atherosclerosis | 0.78 |
|  | Hypertension, Diabetes, and Chronic Kidney Disease | 0.62 |
|  | Hypertension and Diabetes | 0.61 |
|  | Hypertension and Chronic Kidney Disease | 0.60 |
| Caquetá | Hypertension and Diabetes | 0.80 |
|  | Hypertension and COPD | 0.71 |
|  | Malignant Breast Tumor, Obesity, and Hypertension | 0.51 |
| Cauca | Hypertension and COPD | 0.71 |
|  | Hypertension and Diabetes | 0.65 |
|  | HIV/AIDS, Tuberculosis, and Sequelae | 0.64 |
|  | Hypertension, Diabetes, and Chronic Kidney Disease | 0.63 |
| Cesar | Hypertension and COPD | 0.72 |
|  | COPD and Chronic Periodontal Diseases | 0.70 |
|  | Hypertension and Malignant Lung Tumors | 0.65 |
| Córdoba | Hypertension and Diabetes | 0.72 |
|  | Hypertension and COPD | 0.65 |
|  | Chronic Viral Hepatitis and Nutritional Anemias | 0.64 |
|  | Hypertension and Lymphoma | 0.52 |
| Cundinamarca | Hypertension and Prostate Diseases | 0.75 |
|  | Hypertension and Diabetes | 0.72 |
|  | COPD and Chronic Periodontal Diseases | 0.71 |
|  | Hypertension and Chronic Kidney Disease | 0.65 |
|  | Hypertension, Diabetes, and Chronic Kidney Disease | 0.62 |
| Chocó | Hypertension, Diabetes, and Chronic Kidney Disease | 0.78 |
|  | Hypertension and Prostate Diseases | 0.75 |
|  | Hypertension and Diabetes | 0.71 |
|  | Tuberculosis and Diabetes | 0.56 |
| Huila | Hypertension and Diabetes | 0.62 |
|  | Hypertension, Diabetes, and Chronic Kidney Disease | 0.58 |
|  | Hypertension and Atherosclerosis | 0.71 |
| La Guajira | Hypertension and Atherosclerosis | 0.79 |
|  | Hypertension, Diabetes, and Chronic Kidney Disease | 0.78 |
|  | Hypertension and Diabetes | 0.71 |
| Magdalena | Hypertension and Diabetes | 0.82 |
|  | Hypertension and COPD | 0.66 |
|  | HIV/AIDS, Tuberculosis, and Sequelae | 0.64 |
| Meta | Hypertension, Diabetes, and Chronic Kidney Disease | 0.74 |
|  | Hypertension and Diabetes | 0.71 |
|  | Hypertension and COPD | 0.66 |
| Nariño | Hypertension, Diabetes, and Chronic Kidney Disease | 0.74 |
|  | Hypertension and Diabetes | 0.71 |
|  | Hypertension and COPD | 0.66 |
|  | HIV/AIDS, Tuberculosis, and Sequelae | 0.64 |
| Norte de Santander | Hypertension and COPD | 0.76 |
|  | Prostate Cancer and Hypertension | 0.74 |
|  | Hypertension and Diabetes | 0.71 |
|  | HIV/AIDS, Tuberculosis, and Sequelae | 0.64 |
| Quindío | Hypertension and Atherosclerosis | 0.78 |
|  | Hypertension, Diabetes, and Chronic Kidney Disease | 0.72 |
|  | Hypertension and Diabetes | 0.71 |
|  | Hypertension and Chronic Kidney Disease | 0.69 |
|  | HIV/AIDS, Tuberculosis, and Sequelae | 0.64 |
| Risaralda | Hypertension and Atherosclerosis | 0.78 |
|  | Prostate Cancer and Hypertension | 0.74 |
|  | Hypertension and Diabetes | 0.71 |
|  | Thyroid Cancer, Obesity, and Diabetes | 0.65 |
|  | HIV/AIDS, Tuberculosis, and Sequelae | 0.64 |
| Santander | Hypertension and Atherosclerosis | 0.78 |
|  | Hypertension and COPD | 0.76 |
|  | Hypertension and Diabetes | 0.71 |
| Sucre | Hypertension and Diabetes | 0.78 |
|  | Hypertension and Atherosclerosis | 0.71 |
|  | Hypertension and COPD | 0.70 |
|  | Hypertension and Lymphoma | 0.52 |
| Tolima | Hypertension and Diabetes | 0.75 |
|  | Hypertension and COPD | 0.72 |
|  | Hypertension and Atherosclerosis | 0.71 |
| Valle del Cauca | Hypertension and Diabetes | 0.82 |
|  | Hypertension and Prostate Diseases | 0.75 |
|  | Hypertension, Diabetes, and Chronic Kidney Disease | 0.71 |
|  | Thyroid Cancer, Obesity, and Diabetes | 0.65 |
|  | HIV/AIDS, Tuberculosis, and Sequelae | 0.64 |
|  | Leukaemia and Substance Use Disorders | 0.49 |
| Arauca | Hypertension and Diabetes | 0.82 |
|  | Hypertension, Diabetes, and Chronic Kidney Disease | 0.71 |
| Casanare | Hypertension and Diabetes | 0.75 |
|  | Hypertension, Diabetes, and Chronic Kidney Disease | 0.74 |
|  | Hypertension and COPD | 0.76 |
| Caquetá | Hypertension and COPD | 0.76 |
|  | Hypertension and Diabetes | 0.65 |
|  | Cervical Cancer and Depression | 0.52 |
| Putumayo | Hypertension and COPD | 0.76 |
|  | Cervical Cancer and Depression | 0.52 |
|  | Malignant Breast Tumor, Obesity, and Hypertension | 0.51 |
| San Andrés | Thyroid Cancer, Obesity, and Diabetes | 0.65 |
|  | Hypertension and Lymphoma | 0.52 |
| Amazonas | Prostate Cancer and Hypertension | 0.74 |
|  | Malignant Breast Tumor, Obesity, and Hypertension | 0.51 |
| Guainía | Hypertension and COPD | 0.76 |
|  | Hypertension and Diabetes | 0.75 |
|  | Hypertension, Diabetes, and Chronic Kidney Disease | 0.74 |
|  | Cervical Cancer and Depression | 0.52 |
|  | Malignant Breast Tumor, Obesity, and Hypertension | 0.51 |
| Guaviare | Hypertension and Diabetes | 0.75 |
|  | Malignant Breast Tumor, Obesity, and Hypertension | 0.51 |
|  | Chronic Viral Hepatitis and Malignant Liver Tumors | 0.50 |
| Vaupés | Hypertension and Diabetes | 0.81 |
|  | Malignant Breast Tumor, Obesity, and Hypertension | 0.51 |
|  | Leukaemia and Substance Use Disorders | 0.49 |
| Vichada | Hypertension, Diabetes, and Chronic Kidney Disease | 0.74 |
|  | Hypertension and Diabetes | 0.71 |
|  | Leukaemia and Substance Use Disorders | 0.49 |

**Table 4:** Prevalence and Impact of Hypertension and Diabetes Cluster During COVID-19 (2019-2023)

| Year | Prevalence (%) | COVID-19 Hospitalizations (per 1000) | COVID-19 Mortality Rate (%) | Vaccination Coverage (%) |
| --- | --- | --- | --- | --- |
| 2019 | 20.5 | - | - | - |
| 2020 | 22.8 | 45 | 15.3 | 12 |
| 2021 | 24.3 | 50 | 14.7 | 65 |
| 2022 | 23.7 | 38 | 12.2 | 78 |
| 2023 | 22.1 | 30 | 10.5 | 82 |

The highest frequency of multimorbidity was found in the Eastern region with an average of 53.0%, followed by the Coffee Belt region with 45.3%.

### MIPRES Analysis for 2023 in Patients with Multimorbidity

In 2023, MIPRES reviewed prescriptions from 32 EPSs in the subsidized regime (94.0%) and 16 EPSs in the contributory regime (88.0%). Nutritional supplements had the highest non-benefit plan prescriptions, totaling 294,624 in 2017. Antioquia, the Coffee Belt, and Bucaramanga’s metropolitan area had the highest per-affiliate prescriptions, with lower numbers in Bogotá, the Caribbean region, and the southwestern part of Colombia.

The most common cluster, hypertension and diabetes, affected an average of 2,464,885 patients annually, with 2,249,504 prescriptions from 8,611 professionals. Departments like Antioquia, Atlántico, Boyacá, Santander, Risaralda, Quindío, Manizales, Cauca, Caldas, Huila, and Valle del Cauca had the highest occurrence of this cluster with non-benefit plan medications.

### Analysis of a Multimorbidity Cluster Impacted by COVID-19

One of the most significant clusters identified in the study was the combination of hypertension and diabetes, which has been notably impacted by the COVID-19 pandemic. Individuals with these conditions are at a higher risk of severe COVID-19, leading to complications such as acute respiratory distress syndrome, multi-organ failure, and increased mortality rates. Additionally, the pandemic has significantly impacted mental health, causing increased stress, anxiety, and depression, which can adversely affect blood pressure and glucose control.

Stress-related behaviors such as poor diet, lack of exercise, and increased alcohol consumption can further exacerbate these conditions. The data showed that the prevalence of the hypertension and diabetes cluster was particularly high among older adults, who are also more susceptible to severe COVID-19 outcomes. The pandemic has likely increased the burden on this population due to direct health impacts and indirect consequences through healthcare disruptions and socioeconomic stressors (10, 12).

The impact of COVID-19 on the multimorbidity cluster of hypertension and diabetes was analyzed using Structural Equation Modeling (SEM). This analysis was conducted to compare the prevalence, healthcare utilization, and outcomes for this cluster before and after the onset of the COVID-19 pandemic.

### Situation of COVID-19 (2012-2019)

Before COVID-19 (2012-2019), Colombia’s hypertension and diabetes cluster prevalence was 20.5%. Path coefficients were 0.65 for hypertension and 0.55 for diabetes regarding age, and 0.45 for hypertension and 0.50 for diabetes regarding healthcare utilization. Patients averaged 6.8 annual healthcare visits, a 78% medication adherence rate, and a 25 per 1000 hospitalization rate for complications (13).

After COVID-19 (2020-2023), the hypertension and diabetes cluster peaked at 24.3% in 2021 and declined to 22.1% by 2023. Path coefficients rose to 0.68 for hypertension and 0.58 for diabetes concerning age, and 0.55 and 0.60 for healthcare utilization, respectively. The pandemic increased the burden on older adults, with healthcare utilization dropping to 5.4 visits annually in 2020 and rebounding to 6.2 by 2023. Medication adherence fell to 65% in 2020, returning to 78% by 2023. Hospitalization rates for complications rose to 38 per 1000 patients in 2020 and decreased to 30 per 1000 by 2023.

### The SEM analysis reveals several key findings

The hypertension and diabetes cluster prevalence rose during COVID-19, likely due to exacerbated conditions or improved diagnosis. Healthcare visits and medication adherence dropped significantly in 2020 due to lockdowns, system overload, and patient fears. By 2023, healthcare utilization returned to pre-pandemic levels, indicating recovery in healthcare access and delivery.

Hospitalization rates for hypertension and diabetes complications surged in 2020, underscoring heightened patient vulnerability during the pandemic. Subsequent declines suggest improved condition management and the impact of vaccination and better COVID-19 treatments.

### Limitations and Biases

This study has limitations: reliance on secondary administrative data may cause misclassification bias; its cross-sectional design limits causal inference; potential selection bias in the RIPS database may omit some health interactions, especially among those with limited healthcare access; and overrepresentation of contributory regime patients in the UPC database may affect findings. (14,15).

Residual confounding from unmeasured variables like lifestyle, genetics, and environmental exposures may affect observed associations. The findings’ generalisability is limited by differences in healthcare systems, demographics, and disease prevalence, requiring further research in diverse settings and populations. (16)

## Conclusions

Chronic diseases and their associations are rising in Colombia due to demographic transitions, improved living conditions, new health technologies, and better acute disease treatments, leading to lower infant mortality and increased life expectancy. Multimorbidity is more common in women and those in the contributory regime, likely due to better health-seeking behaviors, treatment adherence, healthcare access, and higher socioeconomic status.

Using the WHO’s definition of multimorbidity, this study classified chronic diseases into 123 groups using ICD-10 diagnoses. Analytical methods included Cluster Analysis, SEM, and Latent Class Analysis across multiple databases. Findings showed hypertension as the most frequent chronic condition, aligning with global trends.

The prevalence of hypertension and diabetes increased notably during the COVID-19 pandemic, attributed to higher risks for severe outcomes and improved diagnosis and reporting. The SEM analysis highlighted significant changes in multimorbidity patterns due to the pandemic, underscoring the need for resilient healthcare systems that can adapt to crises while maintaining essential chronic disease management services. The study emphasized the importance of standardized research approaches to improve comparability and identified hypertension as a critical public health issue. (16, 17)

The study highlights a high multimorbidity prevalence, impacting health service planning and management. It identifies key chronic disease clusters needing targeted interventions for improved health outcomes and reduced healthcare system burden. Continued research and policy development are crucial to tackle Colombia’s chronic disease challenges effectively.

## Data Availability

All data used for the production of the research are taken from public data sources, do not require prior authorization for their use and are freely available. Data taken from: https://www.sispro.gov.co/Pages/Home.aspx

